# e- Health for Fighting Digital Addiction: a Literature Review

**DOI:** 10.1101/2023.12.21.23300213

**Authors:** Artika Nurrahima, Moses Glorino Rumambo Pandin, Ah. Yusuf

## Abstract

**Background:** The internet highly used worldwide triggers addiction to technology at various age stages. Digital addiction has various negative impacts on health, social, economic, and educational aspects. Various recent studies explained the influence of technology-based interventions to prevent and overcome digital addiction, which indirectly reduced the negative impact of addiction. The purpose of this study was to analyze the use of e-health in various age ranges, the type of e-health used and the type of intervention that uses e-health as a media for fighting digital addiction.

**Method:** This study is a literature review related to e-health intervention in digital addiction obtained from data bases: SpringerLink, ScienceDirect, ProQues, Ebsco CINAHL, and Google scholar with keywords and booleans: “internet addiction” OR “smartphone addiction” OR “game addiction” AND therapy OR intervention AND “e-health” OR “digital health” OR “digital intervention” OR “digital education. Articles in this review are limited to English articles from 2020-2023. The PRISMA guidline is used as a reference protocol and evaluation of literature review.

**Result:** Based on the 11 articles included in this review, e-health interventions are used as strategy to prevent and treat digital addiction in children, adolescents and adults. The majority of the population who use e-health are adolescents. The type of e-health used as the main intervention was smartphone apps, while game was used as additional intervention. The majority of interventions that can be delivered through e-health are CBT based, while other interventions are behavior monitoring and nudge based.

**Conclusion:** E-health intervention can be used as the main and additional intervention to prevent and overcome digital addiction in various ages, especially adolescents, tend to have a high success rate. The development of e-health can be seen as a promising intervention to facilitate healthy digital behavior change.

## INTRODUCTION

Technological advances make the internet familiar to use in everyday life. Internet users worldwide as of October 2023 are recorded at 5.3 billion or 65.7% of the total world population. The Asian continent has the largest number of internet users in the world at 2.93 billion, followed by the European continent of approximately 750 million internet users (Petrosyan, 2023). This condition leads to technology addiction or digital addiction.

Digital addiction (DA) DA is the failure to resist an impulsive act or behavior that causes compulsive use of digital, despite the negative impact of digital technology use (Singh & Singh, 2019). Digital addiction occurs in various age groups. Child and adolescent tend to became addicted to electronical devices, online game and social media (Ding & Li, 2023),(Putra et al., 2023), (Macur & Pontes, 2021), (Baciu, 2020). 19-24 years old was population with a great number problem gamers (André et al., 2020).

Digital addiction has various negative impacts. DA has several negative impacts on health, academic, social, and economic achievement. The adverse health impacts experienced by people with internet addiction, including online gamers include: physical, mental health and unhealthy behaviors. Physical health decline in the form of: obesity, neck and spine pain, bone and joint problems, visual impairment and hearing loss (Aziz et al., 2021). Children and adolescents with game addiction show symptoms that lead to mental health disorders such as fatigue, depression and inability to concentrate (Alrahili et al., 2023). Unhealthy behavior due to game addiction can be in the form of sleep pattern disturbances (Zaman et al., 2022), (Baciu, 2020) as well as time changes in activities or sports which then affect physical and mental health (M. Zhou et al., 2022),(Baciu, 2020). The negative effects of game addiction on academics in the form of decreased school performance; social effects in the form of self-isolation and limited interaction with others; Economic effects are economic changes due to money spent on the benefit of playing online games (Abdou et al., 2021).

Various negative impacts of digital addiction, make this problem important to overcome. Current e-health interventions tend to be used as a media to prevent or overcome digital addiction problems, along with the increas used of intermet and electronic devices that utilize the internet. Research that shows the effectiveness of type e health and the type of intervention that can be used through e heath is very limited and needs further investigation, at what stage of human development that e health can be implemented. Therefore, to answer the existing research gap, this review will discuss about: what type of e health is appropriate to use according to the stage of human development and the type of intervention that can be implemented through e health.

## METHODS

This literature review examined the results of research related to e-health intervention in digital addiction. The articles reviewed come from data bases: ScienceDirect, SpringerLink, Ebsco CINAHL, ProQuest and Google scholar. The article search in this review was used keywords and booleans: “internet addiction” OR “smartphone addiction” OR “game addiction” AND “e-health” OR “digital health” OR “digital intervention” OR “digital education” AND intervention OR therapy. Articles in this review was limited to English articles that published in the last 3 years (2020-2023). This study used the PRISMA flow-chart for selecting and recording articles. The articles that met the inclusion criteria: population of digital addicted people with e- health intervention, were included in this study. The articles were excluded using the exclusion criteria: non-digital addicted population, non-e- health intervention, did not discussed digital addiction, review articles (systematic review, scoping review, narrative review, literature review, etc), not used English language, and not full text article. The processes of identification, screening, and exclusion are ilustrated in the PRISMA flow chart below (Figure 1).

**Figure 1.**
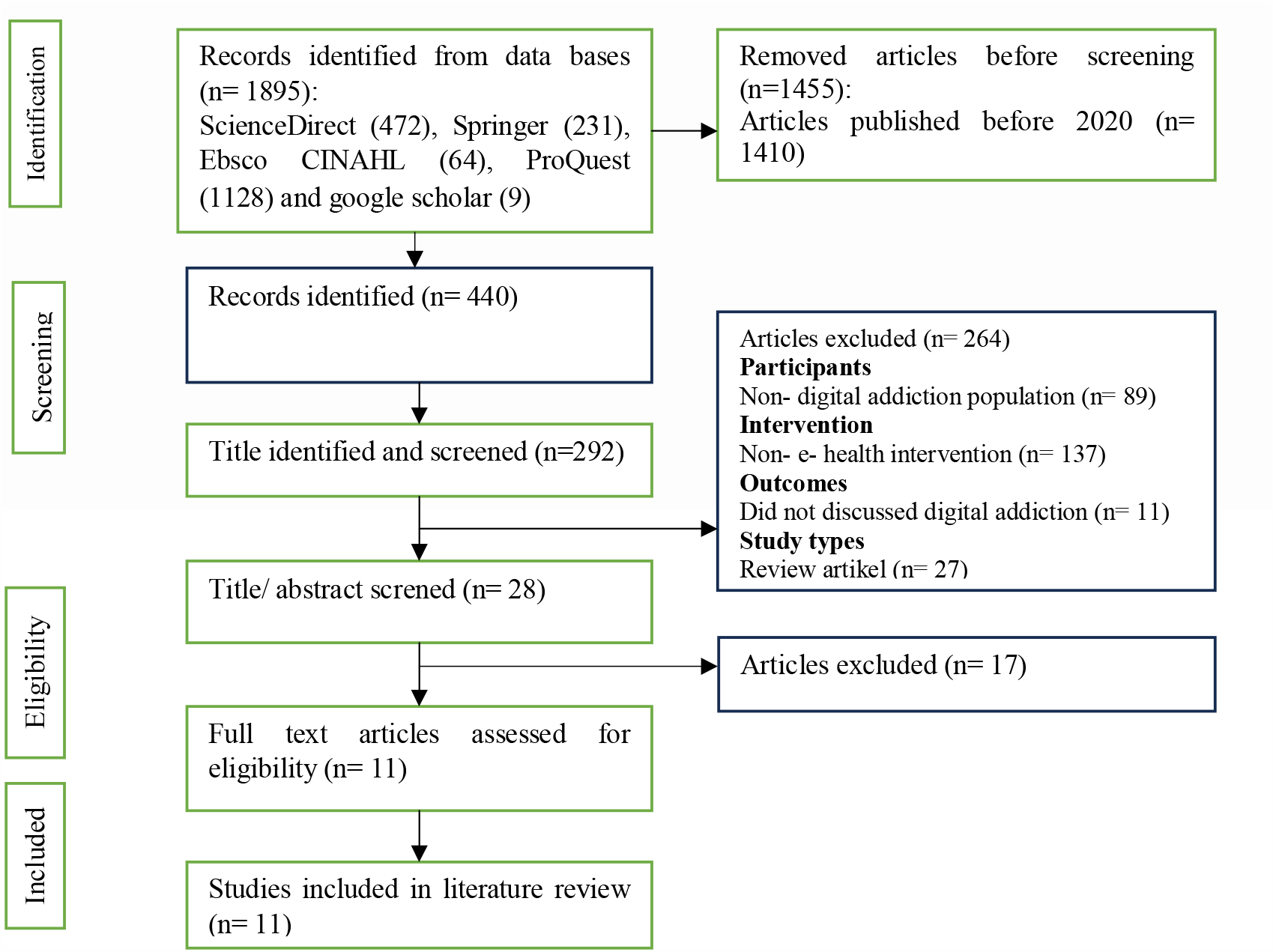
PRISMA flow chart

**Tabel 1.**
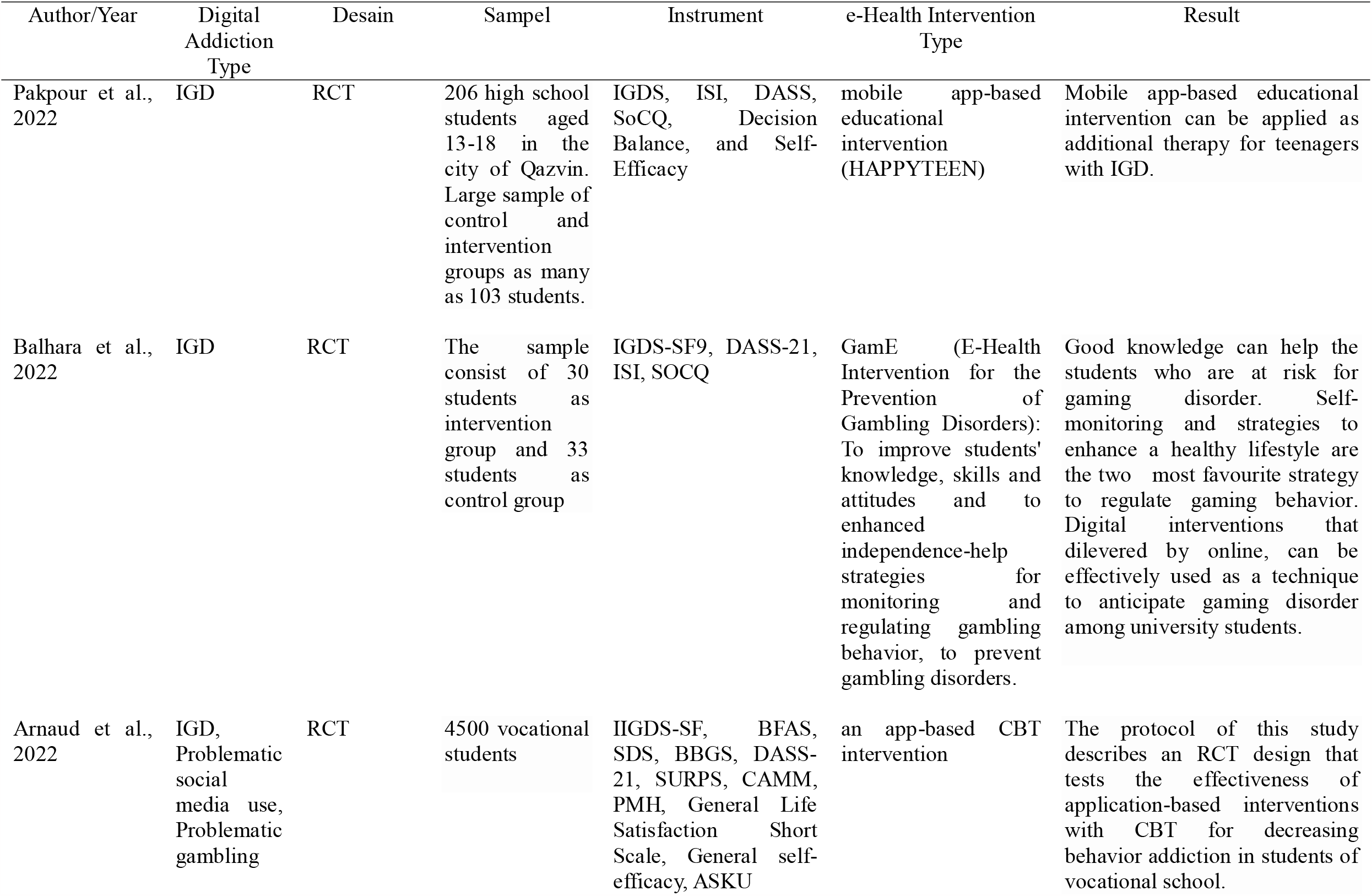
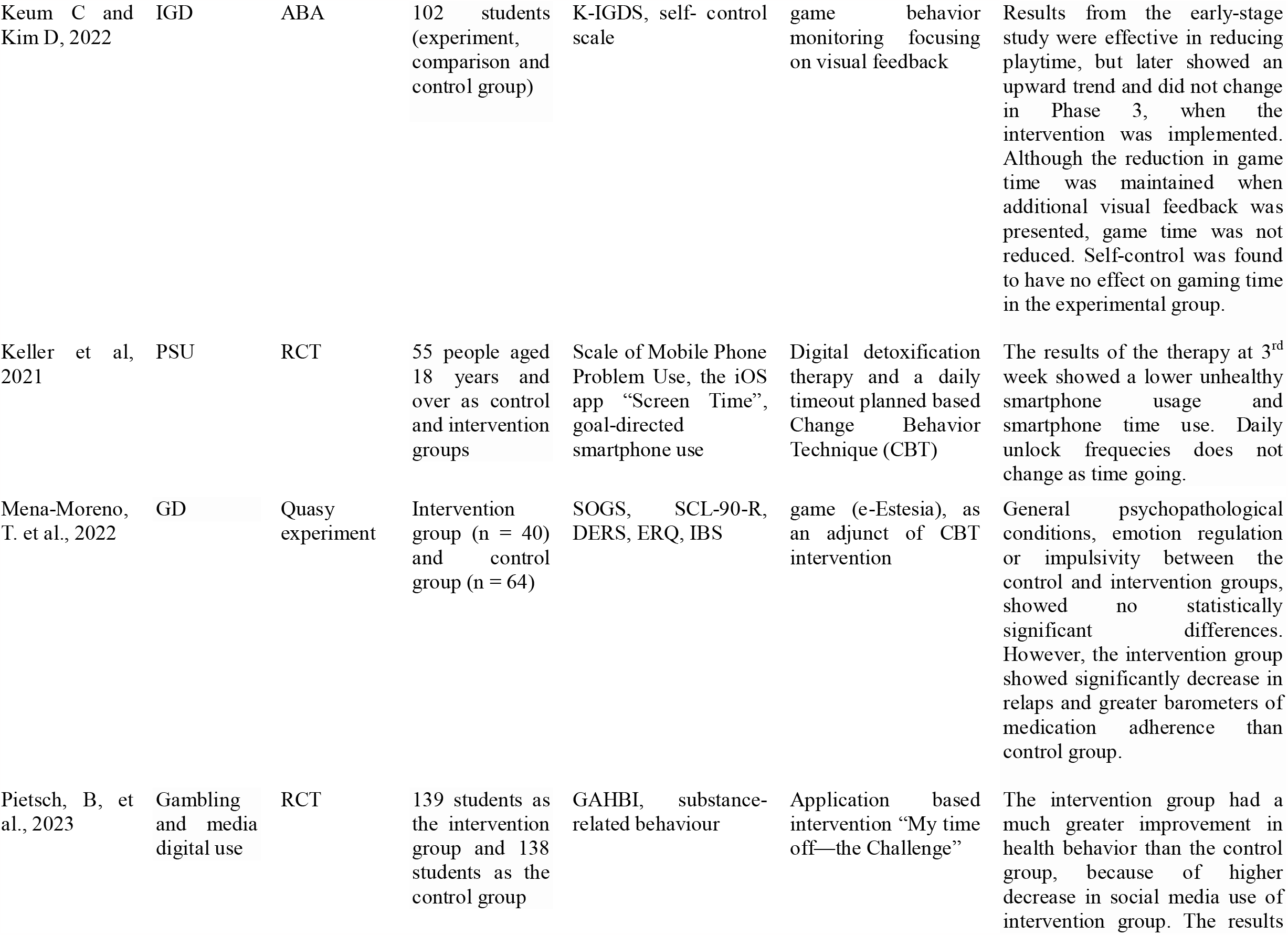
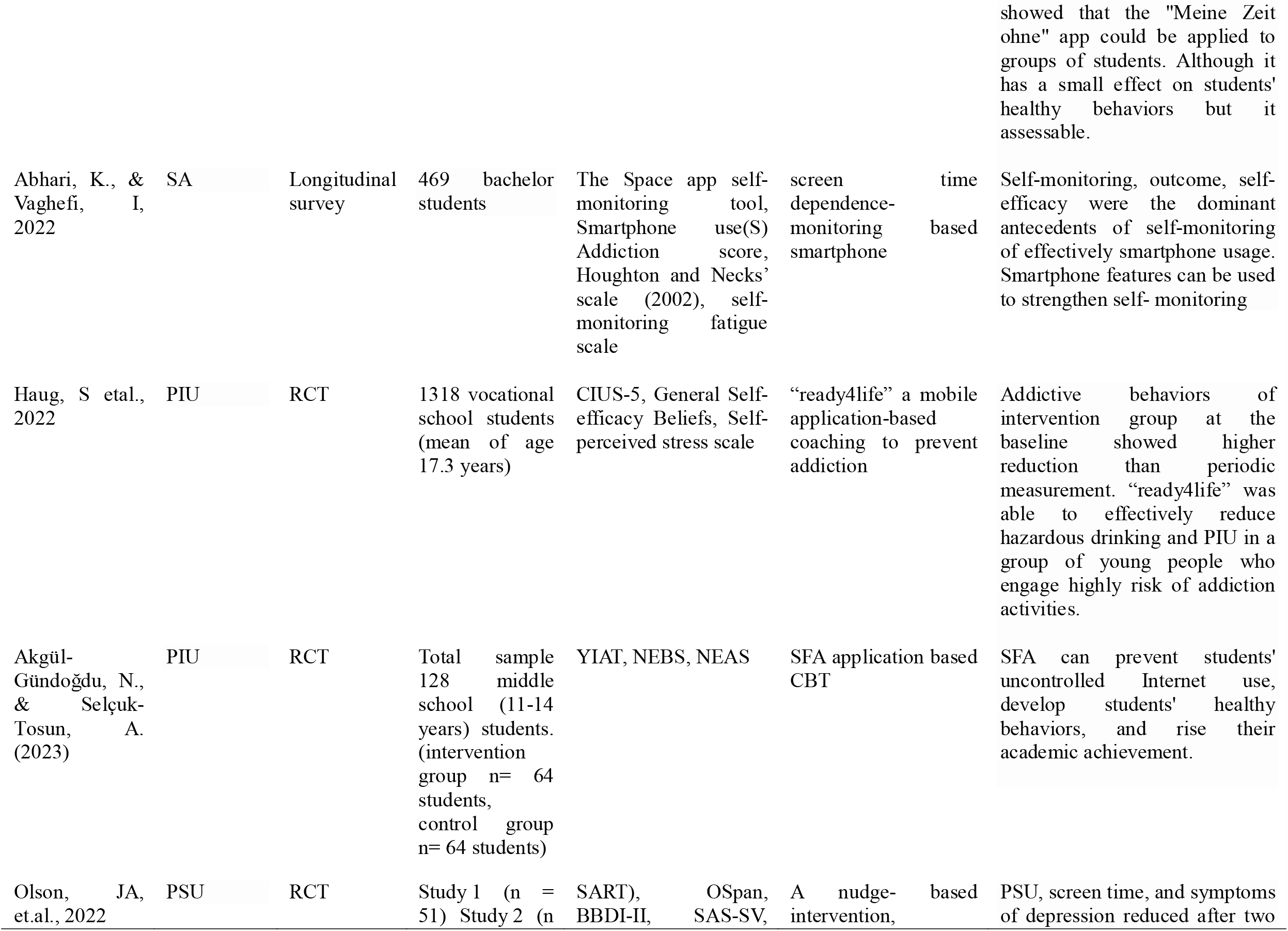
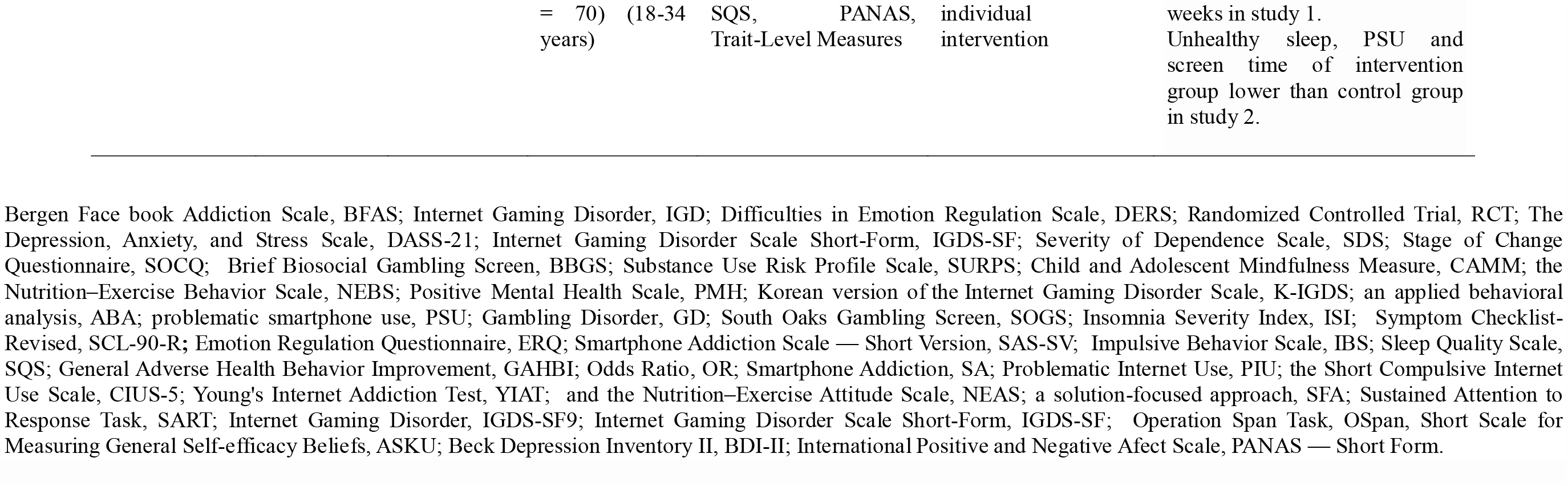
Summary of Study Characteristics.

## RESULT AND DISCUSSION

### The Use of e- health in various stages of human development

E- health as an intervention media that adapts to current technological developments, can be used as an intervention media to overcome internet addiction problems. E-health can be applied in various stages of human development. Digital based intervention is a promising alterative intervention for adolescents with digital addiction (Ding & Li, 2023). Not only in adolescents, e health interventions can also be applied to children starting at the age of 11 years (Akgül-Gündoğdu & Selçuk-Tosun, 2023) and adults aged 18 years and over (Keller et al., 2021). Based on the results of the review, the majority of e-health is applied to students, starting from elementary, junior high school students to college students. The digital world is very familiar with student life. Students use the internet and other digital devices for various purposes such as: education (Hidayat et al., 2022), (Suparmi et al., 2020), (Wang et al., 2023); entertainment (Suparmi et al., 2020) and information (Suparmi et al., 2020). Internet and smartphone users among children and adolescents are quite high compared to other age stages. The internet use of 15-24 years old people was high worldwide. The average of the internet usage globally within 15-24 age population was 75% (Petrosyan, 2023). This is because, adolescents are more adaptable with technological development (Gezgin et al., 2018).

### e- Health Types

Two types of e-health to overcome digital addiction that has been done in previous studies are: mobile apps and games.

### Mobile app

Mobile app is a popular application used today. As many as 6 of the 11 studies reviewed in this article use mobile apps. First, an educational application called HAPPYTEEN based on TTM theory and CBT is used as e health to overcome Internet Gaming Disorder (IGD) (Pakpour et al., 2022). In addition, the application also produces secondary outcomes in the form of anxiety, decision balance, depression, insomnia, stages of change, self-efficacy, and stress. Participants are given a special code (email or phone number) to be able to enter the application. This application is acceptable and easy to understand by teenagers. Participant activities in using the application include accessing each page, reading time of every page and login frequencies. This application contains education about the emergency room and interventions to reduce stress and increase self-efficacy such as: relaxation techniques, yoga (audio file), mindfulness training and mental imagery (Pakpour et al., 2022).

Second, My time off—the Challenge” is an app-based intervention designed to use in vocational college (Pietsch et al., 2023), (Arnaud et al., 2022). This application facilitates a voluntary effort to reduce or stop consuming certain substances, gambling and excessive media use for two weeks. This application is an attempt to prevent certain drug use, gambling, or excessive media use in a school environment using the voluntary abstinence paradigm. The students should install an app, then select an important behavior change task from their smartphones. They can quit from smoking, cannabis, alcohol, use of digital media, gambling, or “other habits” for 14 days or reduce their behavior. If students selected the latter option, they were asked to define their abstinence goals. After that, participants will receive daily notifications, automatically sent in the morning and afternoon, to rate how confident they were in maintaining their goals over the next 24 hours. They were also asked if they had achieved their goals the earlier day. Students were allowed to download and share their certificate of completion, no matter what is the outcome of the assignment on the 14^th^ day (Pietsch et al., 2023), (Arnaud et al., 2022).

Third, Space app and free screen time monitoring app application facilitates successful screen time independent-monitoring grounded on self-monitoring action (Abhari & Vaghefi, 2022). Using this application, participants were able to arrange regular use of smartphone, aims (daily, weekly, etc.) and find out usage compare to set goals. The self-monitoring using smartphones increases awareness of smartphone use and leads to positive outcomes for users (Abhari & Vaghefi, 2022).

Fourth, “ready4life” is a mobile application -based preventive addiction program for trainees that fosters life skills and decreases risky behaviors in the school environment (Haug et al., 2022). The mobile app based coaching program provides personalized coaching through conversational agents. The program aims to reduce trainee risk by enhancing life skills (stress management, social skills) and lowering various risk behaviors (problematic internet use, tobacco/vaping). The program was effective in decreasing addictive behaviors, especially dangerous alcohol consumption and PIU (Haug et al., 2022).

Fifth, Solution Focus Approach (SFA) helps students prevent uncontrolled Internet use, develop healthy behaviours, and obtain academic success (Akgül-Gündogdu & Selçuk-Tosun, 2023). SFA is an effective, structured, and short-term, counseling approach. Implementation of the SFA application will take place only after 6 months, focusing on individual problems, focus on solving problems, powers, and remaining resources (Akgül-Gündogdu & Selçuk-Tosun, 2023).

### Game

Game as a media of addiction intervention is challenging. On the one hand, games are one of the addiction media itself. On the other hand, games are a media that attracts internet addicts to be actively involved in efforts to improve addiction behavior. e-Estesia is an app-based serious game designed to decrease excitement, boost emotional control, and elevate feelings of well-being (Mena-Moreno et al., 2022). The game is set on an island, and the player’s perspective is several meters inland and moves along the coast. Each session he lasts 10 minutes. Since e- Estesia participants had significantly lower relapses and repaired adherence indicators compared to the control group, it can be an additional intervention for GD (Mena-Moreno et al., 2022).

### Types of interventions

The types of intervention that can be delivered through e- health was vary. Interventions based on CBT, monitoring, nudge that have the ultimate goal of changing the behavior of people with internet addiction.

### CBT- based

Cognitive behavioral therapy (CBT)- based is a technique commonly used to change behavior. CBT is a type of intervention that has fundamental assumption that mental disorders and psychological stress are kept by cognitive factors (Hofmann et al., 2012). The word “cognitive” in CBT means that the intervention focuses on cognitive processes targeting cognition, emotional experience, and behavior. Cognitive processes influence emotions and behavior causally and interactively (Hofmann, 2021). The results of the meta-analysis show that CBT can be applied to cases of addiction and substance use (Hofmann et al., 2012). In the case of digital addiction, CBT is applied through various strategies and approaches, including: education, coaching, solution focus approach and behavioral strategies.

### Behavior Monitoring

Behavioral monitoring is the daily observation and recording of specific behaviors over a period of time and the use of this data to create or modify behavior change plans (behavioral monitoring). Behavior Monitoring includes feedback. Feedback in behavioral monitoring directly or indirectly provides information to the monitoring process (Keum & Kim, 2022). Behavioral monitoring enabled by providing additional visualization and textual data may be effectively inducing behavioral change (Keum & Kim, 2022). The experimental group’s play duration, once reduced by monitoring, was kept by visual feedback, while a significant improvement in play time was showed by the comparison group (Keum & Kim, 2022).

### Nudge Based

Nudge-based Nudges are tools for achieving behavior change (Sunstein, 2022). Nudge-based interventions make minor changes to smartphone settings with the goal of reducing smartphone use itself ((Thaler and Sunstein, 2021 in (Olson et al., 2022), such as default settings. Nudges have generally been proven to be effective in controlling immediate behavioral decisions (nudge effects). Olson recommends several touch-based strategies to reduce smartphone use, including: I use: notifications by not activating unimportant notifications, accessibility (none, activating vibration mode, secret, out of sight, etc.) when unuse; unlock; unable Touch ID/Face ID; apply a password; put it to sleep; When you go to bed, mute your phone (turn off vibrations) and keep it out of reach (for example, across the room). Decrease the brightness of phone, apply grayscale, and set the warmness of phone colors. Lock up or delete email applications and social media in folders on your home screen. If you can complete a task on your computer, save it on your computer (social media, web searches, email, etc.). Tell your family, friends, and colleagues that they are unlikely to answer unless you call them directly. Put phone at home when you don’t require it. Overall, try to minimalize the phone use. A combination of strategies can help with problematic phone use (Olson et al., 2022).

## CONCLUSION

e-health can be used as an intervention to address digital addiction at various stages of human development: schoolchildren, adolescents, adults. The application of e-health is mostly applied to adolescents with digital addiction in school settings. Mobile apps are a type of e-health that is widely applied as the main intervention to overcome digital addiction, while the type of e-health in the form of games is implemented as an additional intervention for digital addiction. The types of interventions implemented through e-health media are mostly CBT-based, while other interventions are behavior monitoring based and nudge based.

## Data Availability

All data produced in the present work are contained in the manuscript

